# “Characteristics and stability of sensorimotor activity driven by isolated-muscle group activation in a human with tetraplegia”

**DOI:** 10.1101/2020.06.02.20117036

**Authors:** Robert W. Nickl, Manuel A. Anaya, Tessy M. Thomas, Matthew S. Fifer, Daniel N. Candrea, David P McMullen, Margaret C. Thompson, Luke E. Osborn, William S. Anderson, Brock A. Wester, Francesco V. Tenore, Nathan E. Crone, Gabriela L. Cantarero, Pablo A. Celnik

**Affiliations:** Department of Physical Medicine and Rehabilitation, Johns Hopkins School of Medicine, Baltimore, MD, USA; Department of Biomedical Engineering, Johns Hopkins School of Medicine, Baltimore, MD, USA; Department of Neurology, Johns Hopkins School of Medicine, Baltimore, MD, USA; Department of Neurosurgery, Johns Hopkins School of Medicine, Baltimore, MD, USA; Research and Exploratory Development Department, Johns Hopkins Applied Physics Laboratory, Laurel, MD, USA; National Institute of Mental Health, National Institutes of Health, Bethesda, MD

## Abstract

Understanding cortical movement representations and their stability can shed light on robust brain-machine interface (BMI) approaches to decode these representations without frequent recalibration. Here, we characterize the spatial organization (somatotopy) and stability of the bilateral sensorimotor map of forearm muscles in an incomplete-high spinal-cord injury study participant implanted bilaterally in the primary motor and sensory cortices with Utah microelectrode arrays (MEAs).

We built the map by recording multiunit activity (MUA) and surface electromyography (EMG) as the participant executed (or attempted) contractions of 2 wrist muscles on each side of the body. To assess stability, we repeatedly mapped and compared left--wrist--extensor-related activity throughout several sessions, comparing somatotopy of active electrodes and neural signals both at the within-electrode (multiunit) and cross-electrode (network) levels.

Body maps showed significant activation in motor and sensory cortical electrodes, with fractured, intermixed representations of both intact and paralytic muscles. Within electrodes, firing strength stability decreased with time, with higher stability observed in sensory cortex than in motor, and in the contralateral hemisphere than in the ipsilateral. However, we observed no differences at network level, and no evidence of decoding instabilities for wrist EMG, either across timespans of hours or days, or across recording area. These results demonstrate first-time construction of a bilateral human sensorimotor map with MEAs. Further, while map stability differs between brain area and hemisphere at multiunit/electrode level, these differences are nullified at ensemble level.

## INTRODUCTION

Brain—machine interfaces (BMIs) to restore motor function rely on decoders to infer intended movements from sensorimotor neural activity. Decoder efficacy depends both on the relevance of neural regions sampled---how closely they map to executed (or attempted) movements---and the stability of the regions---how consistent their activity patterns are over time.

Many efforts have been made to topographically map the body onto the primary motor (M1) and sensory cortices (S1)^1-5^. Historically, stimulation studies in M1 indicated that individual body parts are represented with a partially-fractionated somatotopy; namely with the face, arm, and legs seated in largely distinct areas, and individual muscles within an effector represented in a more mixed and overlapping fashion^6-7^. A recent study using microelectrode arrays (MEAs) implanted in a human BMI participant however, provided evidence of intermixed whole-body tuning within the hand knob area^8^, raising questions about M1 organization. The primary somatosensory cortex (S1)^2^, by contrast, exhibits particularly clear somatotopy for cutaneous touch, with proprioception having relatively greater overlap between neighboring regions^9-10^.

Although much work has focused on characterizing the layout of the sensorimotor map, we know less about its stability. One can examine stability in the neural representation of a given movement at multiple levels: as consistencies in the location of active neurons (somatotopy), in the firing patterns at individual recording sites, and in the activation pattern of all electrodes (network level). Studies of neural stability at individual-channel and cross-channel (population) levels have been limited to motor areas and focused mostly on non-human primates (NHPs) without spinal injuries. At the single-channel (or unit) level, there is conflicting evidence about whether firing rates or tuning patterns stay consistent for several days^18-19,23^ or whether they are more typically limited to more modest periods of several minutes to a few hours^20-21, 23-24^.

One may also conceptualize stability as the regularity in covarying activity across multiple recording sites at the ensemble or network level^25-26^. Studies at this resolution have shown that the representation between network-wide activity and reaching movements may be stable through multiple weeks in NHPs^26^, and even a few years in the context of an ecologically important task like targeted reaching^19^. Unfortunately, replications of this approach in humans are limited because of a comparative shortage of studies and the lack of intact, consistently replicable movements to analyze in BMI patients.

Questions of stability are practically relevant. BMIs trained in humans have largely failed to replicate performance in NHPs, often needing to be retrained multiple times throughout sessions.^25,27-35^. Due to potential instability in the sensorimotor map, understanding how consistently this map and its underlying neural activity patterns persist over time may be key to developing longer-lasting decoders. As BMI usage by humans continues to grow, further studies of body map organization and stability are needed, both to understand how implant location affects signal consistency and quality, and to address recent challenges to the principle of somatotopic organization from MEAs implanted in humans.

Here, we studied the body map and its stability in a tetraplegic human (C5/6 incomplete, ASIA B), the first person to be implanted bilaterally with MEAs in the traditional hand area representation of precentral (M1) and postcentral (S1 Area 1) gyri^36^. We first estimated the sensorimotor map associated with EMG-controlled muscle contractions (or attempted motions) in the forearm above and below injury level. Then we characterized stability of neural activity throughout the map over varying time periods for the left wrist extensor muscle (extensor carpi radialis: ECR). We investigated stability from multiple perspectives: in terms of spatial patterning (somatotopy), within electrode-level signaling, and ensemble-level signaling across brain hemispheres and areas. Maps showed significant motor and sensory cortical activity primarily for the upper limb, encompassing both intact and paralytic muscles. At the single-electrode level, we observed higher stability in sensory cortex than motor, and in the contralateral hemisphere than the ipsilateral. In contrast, distinctions found at the unit level were absent at the ensemble level, all the brain regions being equally accurate and stable for decoding muscle activity.

## RESULTS

During each experimental session, we measured neural activity associated with isolated EMG-controlled muscle contractions using a metronome-paced task (Fig. 1; see *Materials and Methods* for details). Given that electrode implants were targeted in the hand knob of motor cortex, and in the hand and fingertip representations in sensory cortex (Fig. 1A), we concentrated this study on wrist and finger muscle contractions (Table 1). Sessions consisted of blocks, in which we instructed the participant to contract (or attempt to contract) a specific muscle in isolation to an audiovisual metronome paced at approximately 1 tick per 4 seconds (Fig. 1B). To assure that contractions were isolated, we trained the participant’s movements under the guidance of physical therapists, and simultaneously monitored electromyograms (EMG) for the instructed muscles and surrounding ones likely to be co-activated. We labeled as noncompliant any trials where EMG was absent, or the participant co-contracted muscles outside of the wrist. We excluded such noncompliant trials from further analysis. As shown in Figure 1C, muscle contractions elicited MUA responses, which we quantified in terms of peri-event time histograms (PETH) computed from thresholded firing rates (see *Materials and Methods*). We performed subsequent analyses on PSTHs with respect to windows relative to the burst onset.

**Figure 1:**
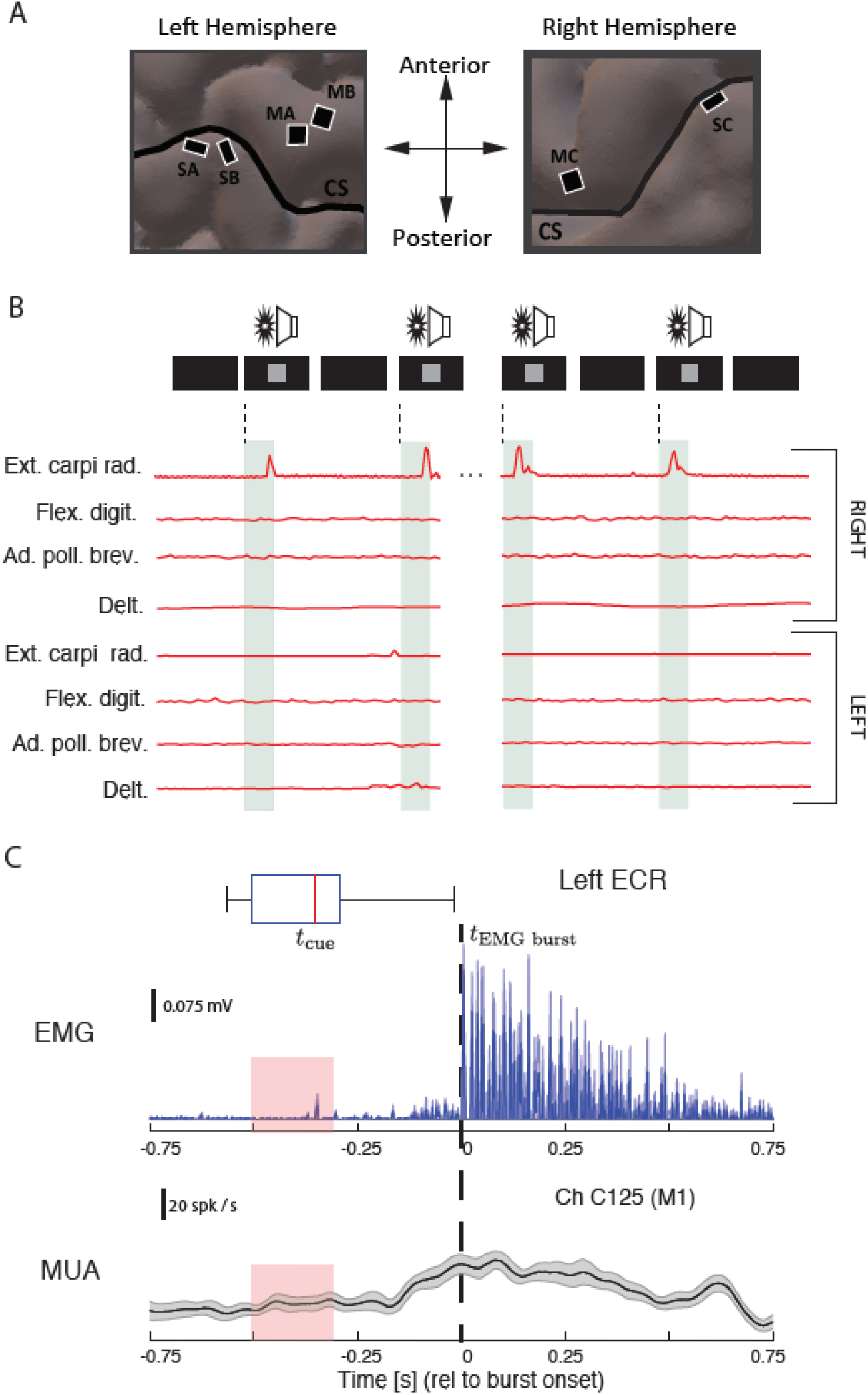
Cortical recording sites and experimental methods. (A) Sites of the six bilaterally-implanted microelectrode arrays (MEAs), overlaid on MRI reconstruction of participant’s brain (CS: central sulcus). Arrays are labeled by anatomical target and pedestal (e.g. MA: primary motor cortex, pedestal A; SB: primary sensory cortex, pedestal B). (B) Experimental paradigm. Isolated muscle contractions (or contraction attempts) were cued by metronome ticks (click and pixel flash) at 4-second intervals plus jitter. Electromyography (EMG) traces are from 4 example trials (repetitions), where the participant executed left extensor carpi radialis (ECR) contractions without co-contracting neighboring or opposite-limb muscles. (C) Temporal referencing and synchronization of neuromuscular and cortical data for a representative EMG-producing muscle (the left extensor carpi radialis: ECR). The upper plot shows event-referenced raw EMG in mV (reference line). Box plot shows movement cue time distributions relative to burst onset; pink regions are the interquartile range (IQR) of this distribution. The lower plot shows the peri-event time histograms (PETH) of multiunit activity (MUA) from an example motor channel on the contralateral motor array, in spikes / sec. Signals were referenced to EMG burst onset (dashed line; see *Supplemental Information*), and are trial averaged. Shaded regions show bootstrapped 95%--confidence intervals.

Overall, we tested contractions of 2 muscles in the wrists (Fig. 2A, B). Figure 2B tabulates, for each muscle, the significantly active channels and their laterality (contralateral, ipsilateral, or bilateral). Overall, 105 channels in M1 and 92 channels in S1 responded to contractions within at least one wrist muscle. Significant activity manifested in all motor and sensory arrays except for MA. Although muscle contractions could be well isolated to one side of the body, channel activity was present in both hemispheres.

**Figure 2:**
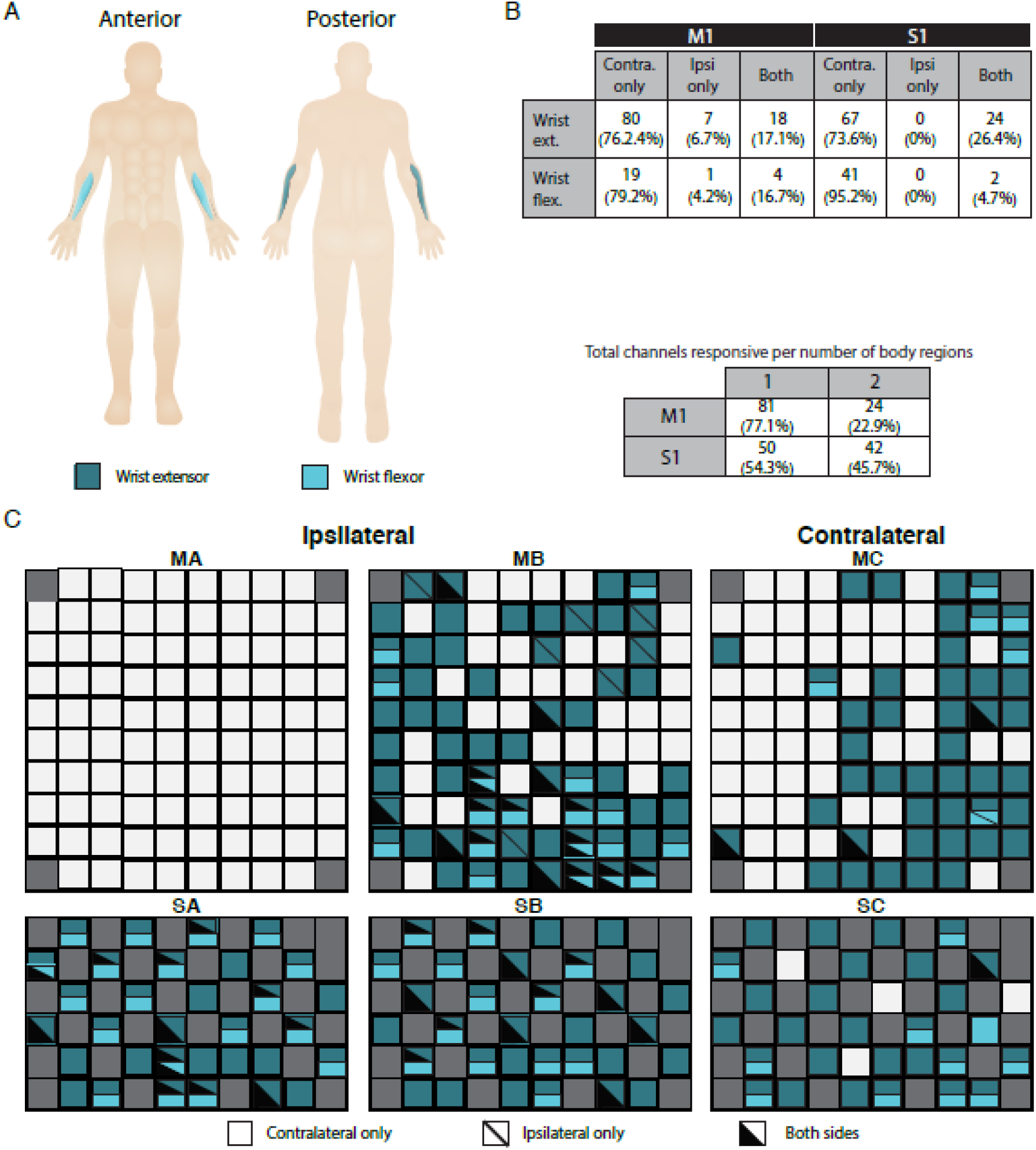
Regional body map for wrists, showing activity patterns across individual muscles. A: Overview of assessed muscles, color-coded by group. Targeted groups were: wrist extensors---extensor carpi radialis; wrist flexors: flexor carpi ulnaris. B: Summary of channel-level representation of each muscle by brain area (M1: motor cortex arrays; S1: sensory cortex arrays) and laterality (contralateral, ipsilateral, or both). Counts are raw numbers, and percentages are with respect to all channels active for that region within the indicated brain area. (C) Activity maps for right hemisphere (MA: motor array, pedestal A; MB: motor array, pedestal B; SA: sensory array, pedestal A; SB: sensory array, pedestal B) and left hemisphere (MC: motor array, pedestal C; SC: sensory array, pedestal C). Channels with significant MUA responses to contractions in a body region are colored as in panel A. Solid colors indicate activity for only the contralateral side of the body (right side for MA/SA/MB/SB, left side for MC/SC); diagonal lines denote activity for the ipsilateral side; and black triangle overlays denote that both sides of the body elicited a response.

In the wrist, unilateral activity accounted for 82.9% of active M1 and 73.6% active S1 channels for the extensor contractions, and 83.3% of active M1 and 95.2% of active S1 channels for flexor contractions. Critically, in a control experimental session where the participant sat quietly listening to the metronome, we found no channels with significant activation on any array.

To perform analyses of map stability, we focused on contractions from the left ECR since the participant could isolate this muscle most consistently. We assessed stability at three levels. First, we examined the consistency and location of active channels across sessions, which we term *longitudinal spatial stability*. Second, within channels, we measured similarities of PETH amplitude (firing strength stability) and PETH shape (firing dynamic stability) over adjacent time periods. Finally, we compared activity patterns across populations of channels, based on the similarity of trajectories on a neural manifold, which we call *ensemble stability*. We provide further details in the *Materials and Methods*, and *Supplementary Information* sections.

### Longitudinal Spatial Stability Declines Faster in the Ipsilateral Hemisphere than the Contralateral, and in Motor Areas than Sensory

We measured longitudinal spatial stability for 12 sessions over a total of 4 months (Fig. 3). We represent longitudinal spatial stability in two ways. First, we plotted *frequency heat maps* of channel activity over MEAs (Fig. 3A). Secondly, we estimated *survival probabilities* for channels on each set of arrays (Fig. 3B-C). Survival probability was the chance that a specific channel was activated for at least *n* total sessions (1 ≤ *n* ≤ 12), not necessarily consecutive. To calculate these probabilities for a given hemisphere or area, we first counted the number of channels in that location that were active for at least *n* sessions. Then, we divided these counts by the total active channels in that location. To describe the shape of survival probability functions, we fit by decaying exponentials of the following form:

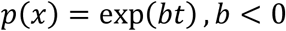

**Figure 3:**
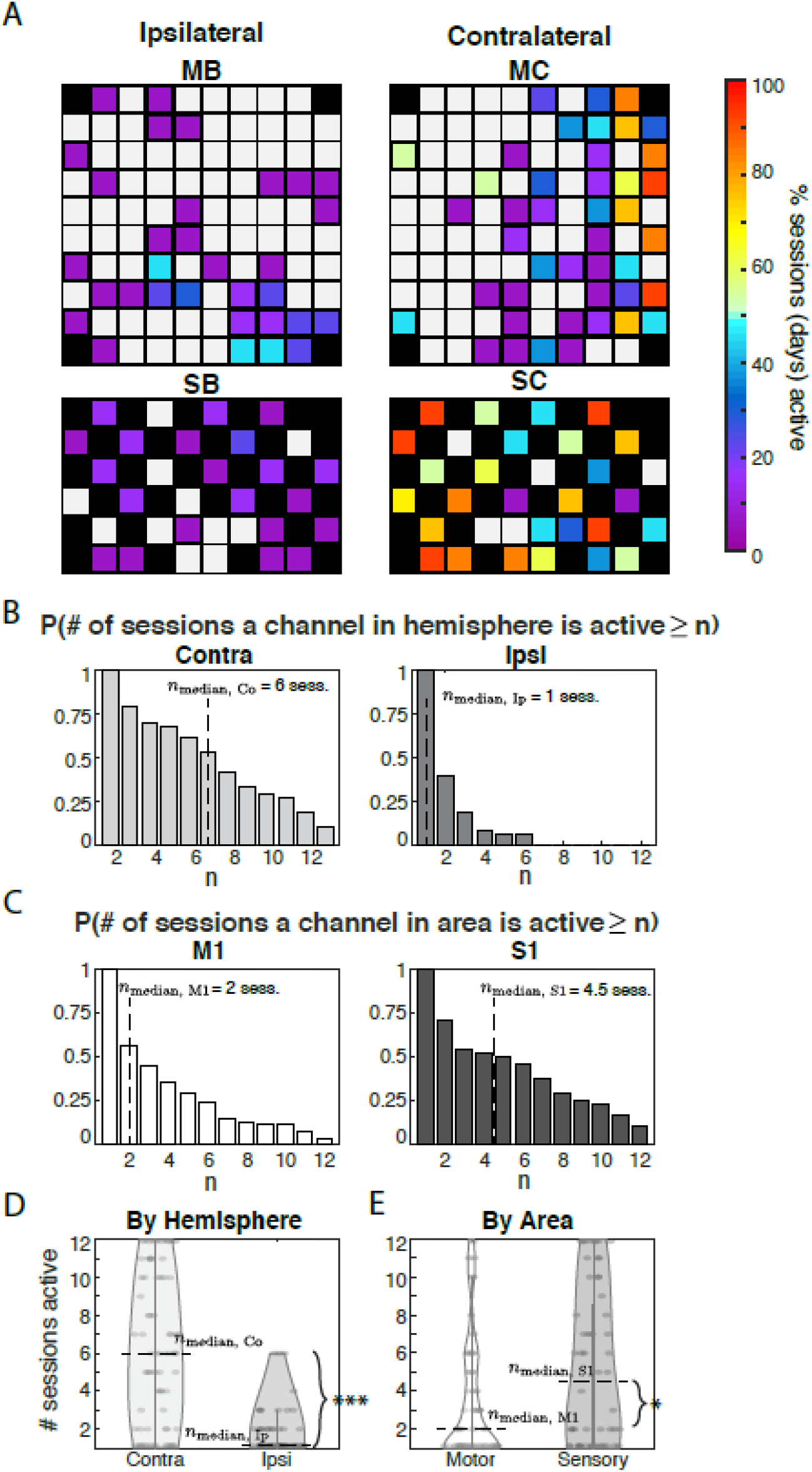
Spatial patterning and longitudinal stability activity from contractions of left wrist extensor (ECR: extensor carpi radialis longus). Longitudinal stability is considered relative to how often a channel is measured as active relative to the number of experiment sessions (12 total). (A) Frequency of activity across sessions for each channel distributed over arrays B and C (top: motor; bottom: sensory). Color code denotes the percentage of sessions (of 12) that a given channel was active, with higher percentages corresponding to greater longitudinal stability. (B) Probabilities that any active channel on Pedestals B and C responds for more than n sessions, within contralateral (left panel) and ipsilateral (right) hemispheres. Motor and sensory areas are pooled. Dashed lines mark the median number of sessions responsive among channels within the active footprint of each hemisphere. (C) Probabilities that any active channel on Pedestals B and C (mapped in panel A) responds for more than n sessions, measured for motor (left panel) and sensory (right) areas (hemispheres pooled). For example, a channel active for exactly 2 out of 12 possible sessions would be counted in the bars for both n=1 and n=2. Dashed lines mark the median number of sessions that a given channel in the active footprint of each area responds. (D) Distributions of the number of total sessions a channel responds to attempted left wrist extensions (among all channels in active footprint), by area. The median number of sessions a channel within the active footprint was observed responsive was greater among sensory than motor arrays (n_Median, M1_ – n_Median, S1_ = -2.5 days, p = 0.022) (E) Distributions of the number of total sessions a channel responds to attempted left wrist extensions, by hemisphere. The median number of sessions a channel was observed responsive was greater within the contralateral than the ipsilateral hemisphere (n_Median, Co_ – n_Median, Ip_ = 5 sessions; p < 0.001)

Arrays on the contralateral hemisphere of the brain (right hemisphere: Pedestal C) had concentrations of highly stable channels (active > 75% of sessions, indicated by warmer colors), which were surrounded by zones of relatively less stability (cooler colors). This pattern was consistent with previous fMRI mapping literature, which resembles a “center of gravity” arrangement. In the motor array, longitudinal spatial stability was highest on the right side of the array (lateral to the brain midline), and declined progressively toward the left (medial) side. In the sensory array, by contrast, the most stable channels were more dispersed, although highest stability tended to lie toward the bottom left (anterolateral) section of the array. The survival probability of contralateral channels across both areas was well fit by an exponential (R^2^=0.938) that appeared to decay at a slow rate (b = -0.127).

In comparison, ipsilateral arrays (Fig. 3A, Pedestal B) showed only sporadic placement of active channels. Activity was largely restricted to < 25% of sessions and exhibited no clear centers of gravity. The survival probability of active ipsilateral channels was also well fit by an exponential (Fig. 3B, right panel; R^2^ = 0.842) that decayed at a sharper rate than in contralateral channels (b = -0.434). Consistent with this difference, the median “lifetime” of contralateral channels was higher than ipsilateral (Fig. 3D; n_Median, Co_ – n_Median, Ip_ = 5 sessions; p < 0.001, Wilcoxon signed-rank test). Altogether, longitudinal spatial stability in contralateral channels was higher than in ipsilateral, and displayed a center of gravity pattern that was absent in ipsilateral arrays.

When channels were grouped by area, decaying exponentials also accurately predicted the probabilities of survival for motor (Fig. 3C, left panel; R^2^= 0.935) and sensory channels (right panel; R^2^=0.979). However, survival chances decayed more sharply in motor (b = -0.248) than in sensory areas (b = -0.724). Accordingly, the median “lifetime” of sensory channels was significantly higher than of motor (Fig. 3D; n_Median, M1_ – n_Median, S1_ = -2.5 days, p = 0.022; Wilcoxon signed-rank test). Thus, sensory channels had higher longitudinal spatial stability than motor channels.

We excluded Pedestal A from analysis because its motor array failed to show any responses to left wrist extensions, consistent with our finding in regional body maps (Fig. 2C and S1C). However, spatial patterning and stability on the sensory array in Pedestal A was qualitatively consistent with our observations on the Pedestal B sensory array.

### Firing Strength Stability Within Channels is Higher in the Contralateral Hemisphere, but Comparable between Sensory and Motor Areas

Next, we compared firing strength stability (amplitude of firing rate over time) within active channels between consecutive hours (within-session), and sessions (across days). To measure firing strength stability for a given channel, we calculated a normalized difference of maximum PETH amplitudes for each hour or session being compared (see *Supplementary Information* for details, especially Fig. S2). Figure 4 presents the average of these stabilities across all channels, by brain hemisphere (Fig. 4A) and area (Fig. 4C).

**Figure 4:**
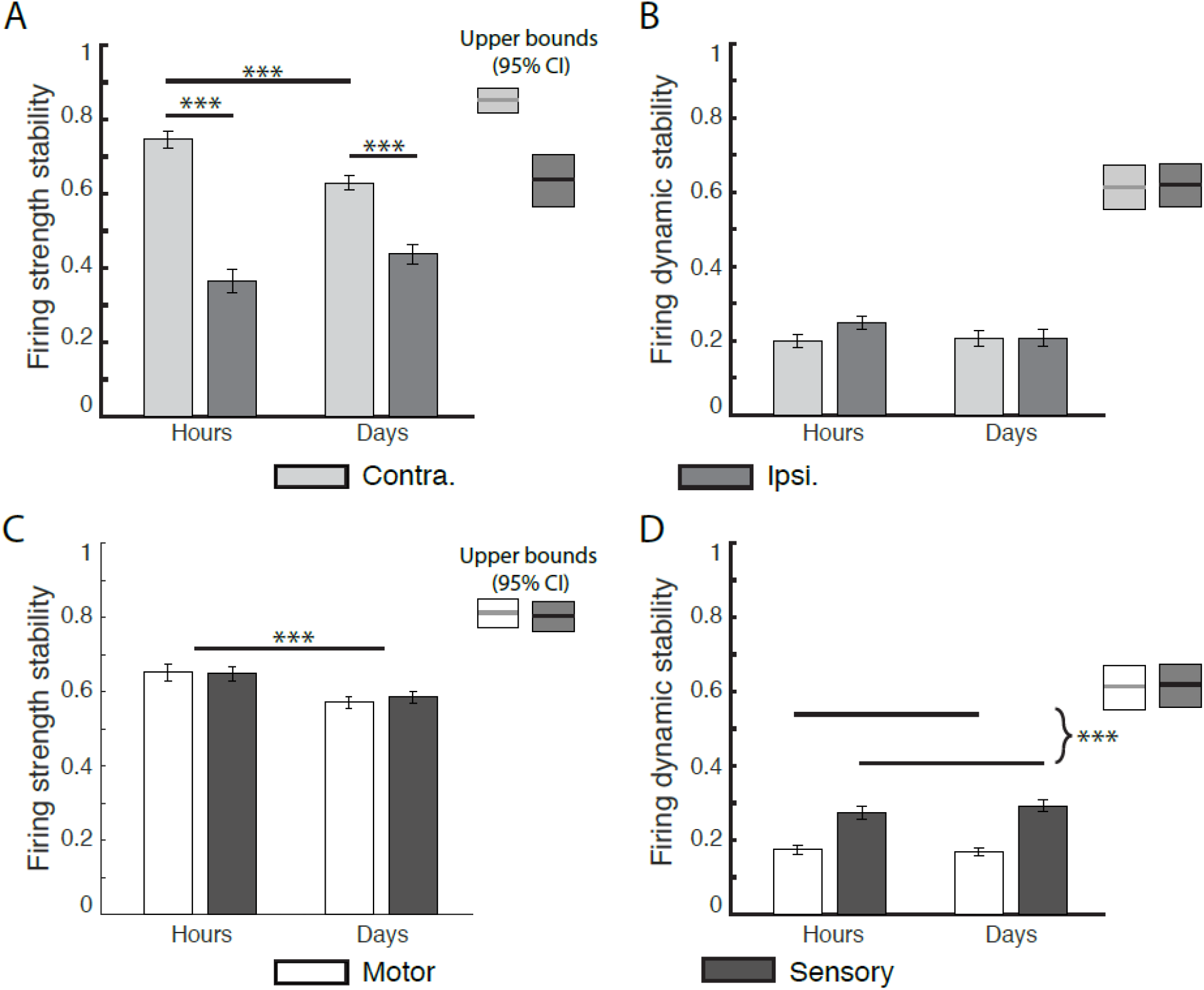
Within-channel (MUA) stabilities of left-ECR-related activity over time. X-axis denotes time scale of comparison (H: hours; D: days). Y-axis measures are normalized to the range [0, 1], higher values denoting greater stability. Bars show mean values +/- 1 SE. (*), (**), and (***) denote significance levels of 0.05, 0.01, and 0.001 respectively.(A-B) Regions labeled Upper bounds are determined by estimating minute-to-minute stability levels by bootstrapping (see *Materials and Methods*). (A) Firing strength stability significantly decreased from hours to days, and was higher for contralateral than ipsilateral channels. There was a significant laterality—by--timescale interaction, with contralateral channel stability decreasing more quickly than ipsilateral. Both hemispheres were less stable within hours than their upper bounds. (B) Firing dynamic stability (cross-correlation between z-scored PETHs) did not significantly decrease from hours to days in both hemispheres. There was a significant laterality—by--timescale interaction, with stability in ipsilateral channels degrading faster than contralateral channels over minutes to hours, such that there was no difference between hemispheres over hours and days. Dynamic stability in both hemispheres was less stable within hours than their upper bounds, suggesting short-term destabilization within minutes to hours. (C-D) Comparative stability across areas, pooled over brain hemisphere. (C) Firing strength stability (relative change in z-score of firing rate) by cortical area, over time. Stability significantly decreased over time from hours to days, with no significant difference between cortical areas. Within-hours stabilities for each area were significantly lower than the within-minutes upper bounds, suggesting an additional decrease on the order of minutes to hours. (D) Firing dynamic stability was invariant within areas from hours to days, but was significantly higher in sensory than motor channels. Within-hours stabilities for each area were significantly lower than the within-minutes upper bounds, suggesting a decrease on the order of minutes to hours.

Comparing firing strength stability between hemispheres (contralateral vs. ipsilateral hemisphere) and across timescales, we observed a main effect of hemisphere (F(1, 1147) = 209.97, p < 0.0001), and a timescale-by-hemisphere interaction (F(1, 1147), p < 0.001; Fig. 4C). Stability was greater in the contralateral hemisphere than the ipsilateral for all time points of comparison (difference within hours = 0.383, p < 0.0001; difference within days = 0.192, p < 0.0001). Similarly, hemispheric stabilities fell below their empirical upper bounds within hours (Contra: hour-to-hour mean = 0.747, 95% CI of upper bound=[0.810, 0.887]; Ipsi hour-to-hour mean = 0.364, 0.95% CI: [0.553, 0.715]). Overall this indicates that as time interval between measurements increased firing strength stability decreased, with contralateral channels being overall more stable than ipsilateral ones.

Comparing firing strength stability between area (motor vs. sensory areas) and timescale, we found no significant effect between motor and sensory areas, but a significant effect for time (F(1,1147) =14.35, p < 0.0001; Fig. 4A) with firing strength stability decreasing from hours to days (mean difference = -0.0718, p =< 0.0001). Within hours, stability in both diminished below their upper bounds as predicted from bootstrapping minute-to-minute data within blocks (see Materials and Methods), both in motor (hour-to-hour mean=0.651, upper bound 95% CI: [0.762, 0.8543]) and sensory areas (hour-to-hour mean = 0.648, upper bound 95% CI: [0.752, 0.848]). Overall this indicates that firing strength stability in each area decreased over time, as early as within hours,

### Firing Dynamic Stability Within Channels is Comparable Across Hemispheres, But Greater in Sensory Areas than Motor

We calculated dynamic stability within channels as a function of cross correlation between PETH waveforms across timescales (see *Supplemental Information* for more details, and Fig. S2 for an example). Figure 4 presents the average of these stabilities across all channels, assorted by brain area (Fig. 4B) or hemisphere (Fig. 4D).

When comparing firing dynamic stability between hemispheres (contralateral vs ipsilateral) and timescale, we found no significant main effects (hemisphere: F(1, 892) = 2.81, p = 0.094; area: F(1,892)=1.29, p = 0.26), nor a hemisphere-by-timescale interaction (F(1, 892) = 2.29, p = 0.13). However, dynamic stability within each hemisphere was lower than its respective theoretical minute-to-minute upper bound (Contra: hour-to-hour mean=0.211, Upper bound 95% CI = [0.573, 0.680]; Ipsi: hour-to-hour mean = 0.126, upper bound 95% CI = [0.522, 0.649]).

Comparing firing dynamic stability between area (motor vs. sensory areas) and timescale, we observed a significant effect for area (F(1, 892) = 64.32, p < 0.0001). Firing dynamic stability was significantly higher for sensory channels versus motor channels (difference of sensory and motor = 0.0974, p<0.001). Dynamic stabilities in both areas were lower than their estimated upper bounds within hours (Motor: hour-to-hour mean = 0.154, motor upper bound 95% CI=[0.552, 0.672]; sensory hour-to-hour mean=0.227, upper bound 95% CI = [0.559, 0.675]).

In sum, firing dynamic stability decreased with longer time intervals between measurements. No differences were observed between hemispheres, while sensory channels were more stable than motor ones.

### Differences in Channel Sortability Do Not Explain Regional Differences in Within-Channel Stability

As noted in a previous study^27^, neural signals defined by multiunit activity are less stable than those containing activity from single units. To rule out whether the dynamic stability differences that we observed between motor and sensory channels were due to differences in separability, we applied a spike sorting algorithm to our individual channel recordings for the left ECR (wave_clus; see *Supplemental Information*, Cluster Analysis for details ^59, 60^). We then stratified our channels by whether they were sortable or not (i.e. whether the algorithm determined one cluster, or multiple clusters), and compared stability metrics across these groupings.

Across all arrays, sensory channels had a significantly higher number of separable units on average than motor channels (M1 avg: 1.171 +/- 0.008 clusters per channel, S1 avg: 1.803 +/- 0.030 clusters per channel; *t*_*8830*_ *=* -28.89; p < 0.001, Fig. S2A). For firing dynamic stability, when we compared brain area (M1, S1) and multiunit separability (single vs multi-cluster units), we found that multi-cluster channels on the whole were less stable than single-cluster, as indicated by their lower cross-correlation values (Fig. S2B). Within single-cluster channels, we found no difference between brain areas (motor 95% CI = [0.390, 0.422], sensory 95% CI = [0.384, 0.426]), however, for multi-cluster groups specifically, motor channels were less stable than sensory (motor 95% CI = [0.183, 0.234], sensory 95% CI = [0.262, 0.321]). Overall this suggests that firing dynamic stability differences between motor and sensory channels are not driven by differences in unit sortability between motor and sensory areas.

### Ensemble Stabilities Are Equal Across Hemisphere and Area

To measure ensemble stability, we calculated neural trajectories across brain hemispheres and areas using principal component analysis (PCA). Visualizing over the first and second principal components (on the “PC1-PC2 plane”; see Fig. 5A-B), we found marked similarity of trajectory shape, orientation, and amplitude between consecutive hours (A), as well as consecutive sessions (B). Expanding the analysis to the top 6 PCs, differences in latent variable trajectories did not significantly change on average, regardless of whether sessions were hours or days apart (C-D), indicating equal stability with time passage. Likewise, trajectory differences were unaffected by brain hemisphere (C) and area (D).

**Figure 5:**
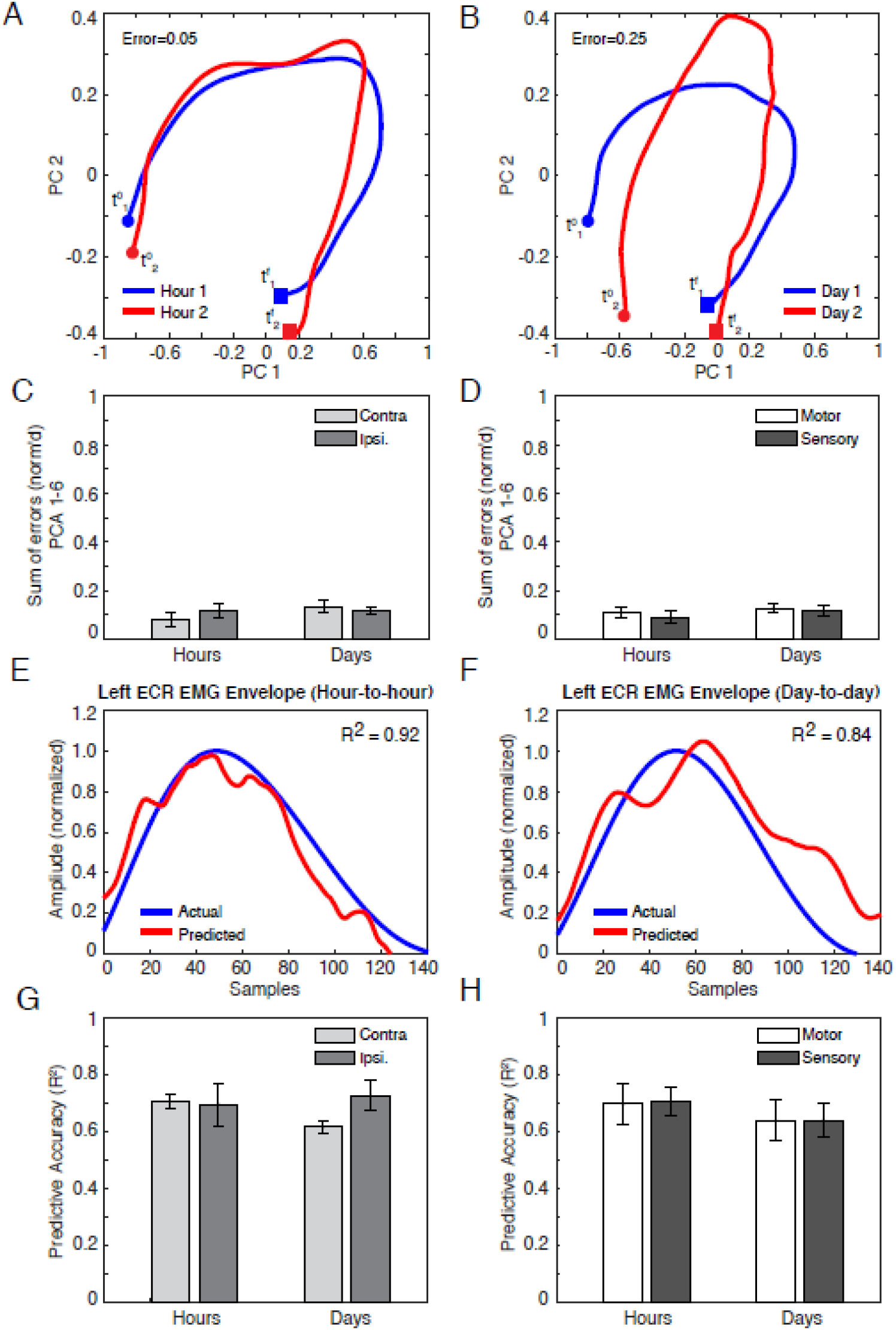
Ensemble-level (network) stabilities of left-ECR-related activity over time. (A–B) Representative principal-component (PC) trajectories of neural ensemble, visualized in the PC1-PC2 plane, during left wrist extensions across typical sets of consecutive--hour (A) and consecutive--day (B) recordings, for the right (contralateral) hemisphere (i.e. motor and sensory channels aggregated). Trajectories reflect average neural activity within EMG bursts only, with endpoints designated at the onset (filled circles, variables t^0^), and terminal points (filled boxes and variables t^f^) of the burst. Error is a normalized Euclidean distance between trajectories, with higher values approaching 1 indicating greater instability (see *Materials and Methods*). (C–D) Cumulative error between trajectory representations of ensemble activity, comprising the first six PCs, between consecutive hours, and consecutive days (Mean +/- 1 s.e). Channel ensembles are grouped by brain hemisphere (C) and area (D). (E–F) EMG of left wrist extensor (ECR: extensor carpi radialis), as measured from surface electrodes (blue) and as predicted from PCA trajectories (PCs 1-6: red) using Wiener filters trained on separate data. Data shown is for test (measurement) and training datasets recorded over consecutive hours (E) and days (sessions) (F). All envelopes are restricted to between burst onsets and terminations, and were time warped to an equal length to facilitate analysis. The x-axis shows the resulting time axis (in samples). The y-axis gives normalized envelope amplitude (see *Supplemental Information*). R^2^ signifies the correlation between the actual EMG and the prediction. (G–H) Correlations (R^2^) between measured (actual) and predicted EMG, arranged by brain hemisphere (G) and area (H). As above, R^2^ is based on training and test sessions spaced over consecutive hours and days (sessions).

Given that such trajectory representations of ensemble activity are of increasing interest for BMI decoders, we then asked how the equivalence in PC stability that we observed over time, area, and hemisphere would translate to decoding muscle activity. To examine decoder stability, we trained a Wiener filter model relating PCA trajectories to muscle EMG envelope (described in *Materials and Methods*, with computational details in *Supplementary Information*). For each pair of data blocks spanning consecutive hours or sessions, we performed a two-fold cross validation procedure. Namely, we trained the Wiener filter on one block, and used the fitted model to predict the envelope of measured left-wrist EMG from the other block. We repeated this process after switching the training and test blocks, and then averaged the goodness-of-fit measures over both cross-validation fits. Depending on whether latent variable inputs derived from motor or sensory inputs, or both, we adjusted terms of model to account for different input latencies (see *Supplementary Information*, and Fig. S4 for neural latency data to verify model assumptions). Typically, there was high correlation between predicted and measured EMG, whether training and test sessions were hours (Fig. 5E; R^2^ = 0.92) or days apart (Fig. 5F; R^2^ = 0.84). Furthermore, on average, we found not only that decoding performance was not significantly affected by the time frame of comparison, nor by area (Fig. 5G) or hemisphere (Fig. 5H). These conclusions did not change when we considered greater numbers of principal components (m = 9 and 12 PCs as in Gallego et al.^26^).

## DISCUSSION

In a study participant affected by a C5/6 incomplete spinal cord injury---the first to be bilaterally implanted with MEAs---we studied how the stability of wrist muscle representation in the sensorimotor cortex across multiple areas, time intervals and scales of resolution. These questions are crucial, in light of practical experiences that human BMIs need to be retrained throughout use sessions. Our main findings were: (1) Channel-level signal stability decreased over time, namely for firing strength; (2) Within-channel stability was higher for sensory channels than motor, particularly in firing dynamics, and higher for contralateral for ipsilateral channels, particularly in firing strength; (3) At the ensemble level, no stability differences were observed in manifold trajectories, either over time or by brain region (area or hemisphere).

### Multiunit Activity destabilizes within short timescales

Intracortical-level topographies of the motor and sensory maps agree with previous findings using non-invasive techniques. In motor areas, we found a concentrated area of maximal stability (see Fig. 3A, array MC) akin to a “Center of Gravity” arrangement^40^ with a surrounding penumbra of less stable channels that disappear and reappear intermittently over sessions^41^. In sensory cortex we found that maximally stable channels were more interspersed, which is consistent with descriptions of S1 somatotopy having multiple dispersed “centers of gravity”^42^.

Our longitudinal mapping of the left-wrist extensor suggests that the somatotopic map may be inherently unstable at the multiunit level, with minimal to no channels being active consistently in all sessions. Within-channel firing strength and dynamic stabilities declined in the order of days, and often sooner within experimental sessions (i.e. hours). Destabilizations of within-channel unit activity within hours and days could be explained by a few factors. For example, MUA signal attributes themselves may have been inconsistent. Because firing rates derive from thresholded activity^43^, our measures are ultimately related to the amplitude of action potential signals, and previous studies have found that action potential amplitudes typically change progressively over hours and days^20,21,23^. Also, MUA instabilities over hours and days could reflect neural plasticity due to our participant’s involvement in BMI activities during the intervening times. Namely, previous work has shown that learning and performance of motor control tasks^26,44^ as well as consolidation during sleep after BMI use^45^ can produce detectable changes in unit-level activity.

### Contralateral MUA is More Stable than Ipsilateral for Longitudinal Spatial Stability and Firing Strength

Sensory and motor channels were more stable in the contralateral than the ipsilateral hemisphere. While the contralateral channels were present during all 12 sessions (Fig. 3B, left panel), the probability of observing any ipsilateral channel for more than 6 sessions was zero (Fig. 3B, right panel). Comparing the median number of sessions that a channel in each area was present, we found that the “lifetime” of an contralateral channel was significantly higher, indicating that ipsilateral representations are inherently more transient. Within sessions, firing strength stability was greater in the contralateral hemisphere than the ipsilateral for all time points of comparison (Fig. 4A). These findings align with a previous fMRI investigation showing the contralateral hemisphere was more consistently active than the ipsilateral representation when executing an arm movement consistently over multiple days^47^.

### Sensory MUA is More Stable than Motor in Longitudinal Spatial Stability and Firing Dynamic Stability

Sensory activity was more stable than motor. Longitudinal spatial stability was higher for sensory arrays, as reflected in a shallower decay in survival probability (Fig. 3C, left panel. Firing dynamic stability was also higher (Fig. 4B). Greater S1 stability does not appear to be related to unit separability (Fig. S2), but rather may be an intrinsic property of region. This seems consistent with our current understanding of the relative functional roles of sensory and motor cortices: sensory processing favors stability and does not require remapping, while motor control necessitates flexibility, particularly in uncertain environments or learning new skills.

The stability discrepancy could be influenced by different material composition of our motor and sensory arrays as well. As we note in our *Materials in Methods* section, motor array electrodes were platinum tipped (Pt), and sensory array electrodes were sputtered—iridium—oxide—film tipped (SIROF). In support of our hypothesis, a previous study observed a tendency of SIROF electrodes to better retain high-amplitude neural signals than Pt^46^, which would likely affect both of our stability measures.

### Ensemble-level Activation is Stable and Overrides Temporal and Regional Differences

At the ensemble level, we found no significant differences in stability across time (Fig. 5A-D) or between regions (Fig. 5C-D), as measured by the errors between Procrustes-aligned PC trajectories.

High channel—level instability may be a natural consequence of the redundancy the nervous system affords in programming movements^24^. Toward this argument, studies in non-human primates (NHPs) show that, despite the replicability of movements such as arm reaches, neural firing patterns at the unit level may often vary quite significantly due to changes in tuning characteristics^20,24^.

Alternatively, it is possible that signal instability may result from limitations in recording hardware^23,27,46^. Latent variable analysis tempers sources of variation by representing neural activity in terms of dimensions (factors) that have optimally high explanatory ability. Despite the underlying sources of the inter-area and inter-hemispheric differences in single-channel stability that we found, PCA suggests that variations in firing rates at the ensemble-level are comparably stable regardless of implant area or hemisphere.

When we utilized these network-level representations to decode EMG from wrist extensions limited to the left ECR, we additionally found that both areas and hemispheres were equally predictive of muscle activation, and that there were no differences in accuracy across time (Fig. 5E-H). This observation is consistent with recent findings in NHPs^26^, although to our knowledge we provide the first evidence in a human.

### Sensorimotor Array Activity Is Widely Distributed Spatially and Temporally

A distinction of our study is that we employed isolated muscle contractions, and validated their compliance with instructions using both guidance from physical therapists and EMG in real time. Our cortical activity map (Fig. 2) showed intermixed muscle representations in motor and sensory arrays, which aligns with previous characterizations. All contractions predominantly elicited activity in the array contralateral to the muscle. However, we detected ipsilateral activity in the motor arrays, and somewhat more bilateral activity in motor and sensory areas (Fig. 2B).

As expected, motor activity preceded sensory activity (Fig. S3). Because our participant’s injury graded B on the ASIA Impairment Scale,^51^ with substantial tactile sensitivity in the hand, much of the sensory response that we observed is likely from afferent sensory feedback. Interestingly, some sensory channel activity preceded typical afferent delays (approx. 30 ms post-EMG burst) or were simultaneous with EMG burst (Fig. S2, pink shaded region).. We conjecture that these early responses may be indirect evidence of efference copy signaling^10^, although further experiments are necessary to verify this claim.

### Implications for BMI

Practical use of BMI technology relies on developing stable decoders. Our findings suggest that decoders that weight neural activity based on the temporal dynamics of specific channels will have limited longevity. This is because the underlying neural code at the individual-multiunit level changes over short timescales, especially in the motor cortex. Although this feature may be biologically beneficial (i.e. the brain encodes motor commands in a flexible manner), it also complicates the current design of neural decoders. This is because a particular combination of neural activity generated by a muscle contraction at one time expectedly leads to changes in neural activity in future iterations of the same contraction. Therefore, a better long-term strategy may be to account for a thorough description of neural activity embodied in the overall dynamics of the response across channels, as this both equalizes stability across brain regions and extends the time frame that it is maintained before declining^48^. Along this line, recent advances in decoding algorithms that consider whole-trial dynamics and activity patterns at a higher level of abstraction rather than individual electrodes^26, 48^ could both provide longer lasting BMI-decoders, as well as overcome stability differences measured across brain regions.

## MATERIALS AND METHODS

This study was conducted under an Investigational Device Exemption (IDE) by the FDA (G170010) and approved by the Johns Hopkins School of Medicine Institutional Review Board (IRB) and NIWC Pacific Human Research Protection Office (HRPO). A study participant with a C5/6 incomplete ASIA Impairment Scale (AIS) B spinal cord injury^51^ (male, 49 years old at time of surgery, 32 years post injury at time of study; right-hand dominant) was implanted bilaterally with six cortical microelectrode arrays (NeuroPort; Blackrock Neurotech, Salt Lake City, UT): 4 in the dominant (left) hemisphere (motor array, pedestal A= MA; motor array, pedestal B= MB; sensory array, pedestal A= SA; sensory array, pedestal B= SB) and 2 in the nondominant (right) hemisphere (motor array, pedestal C= MC; sensory array, pedestal C= SC; Fig. 1A). The participant had residual control of shoulder and wrist muscles and quasi-intact sensation. All motor array electrodes were platinum tipped (Pt), and all sensory array electrodes were sputtered—iridium—oxide—film tipped (SIROF).

### Biosignal Recordings

#### EMG Recordings

During each block, we measured 8 channels of EMG from the targeted muscle, its contralateral homologue, adjacent muscles likely to be co-contracted, and select postural muscles (AMT-8; Bortec Biomedical, Calgary, AB, Canada). We recorded EMG and photodiode data at a 2kHz sampling rate with Blackrock Neural Signal Processors (Blackrock Microsystems, Salt Lake City, UT), and inspected these signals online using the Central software suite (Blackrock Microsystems). We coded all experimental software and analyses in MATLAB (MathWorks, Natick, MA).

#### EMG Screening and Onset Estimation

For each trial, we first epoched EMG signals for all recorded muscles into a window from -0.750 to 2 seconds relative to the metronome cue (see Fig. 1). Then, we visually screened EMG activity for co-contractions, spasms (synchronous high-amplitude global muscle activations), or missed cues (lack of EMG response from an intact target muscle). Afterward, we manually tagged EMG burst onsets and endings using a custom graphical user interface.

#### Neural Recordings

We recorded multiunit activity (MUA) data by automatically thresholding 30-kHz-sampled neural data with Neuroport Neural Signal Processors (Blackrock Neurotech, Salt Lake City, UT), at a level of -3.25 dB relative to quiet sitting. Prior to offline analysis, we binned spiking times into peri-event time histograms (PETHs) at a resolution of 1 ms. Subsequently, we computed all neural activity measures on raw PETHs, only smoothing them for visualization purposes (using Gaussian kernels of width 120 ms) or for ensemble analyses (width 150 ms). We coded all experimental software and analyses in MATLAB.

### Body Map Experiment

We characterized the body sensorimotor map and its underlying MUA firing rate activity within the coverage of the MEA implants for the wrists and hands (Table 1). To evoke muscle activity, we asked the participant to perform (or absent functional movement, to attempt) paced and repeated isolated muscle contractions to a metronome, while simultaneously recording surface EMG and MUA (Fig. 1B). We assessed each muscle in separate blocks of trials. Prior to each block, we made sure the participant was positioned to eliminate postural and extraneous EMG activity outside the targeted muscle. The participant then rehearsed the specified muscle contraction while a trained experimenter monitored EMG activity and provided verbal feedback to facilitate proper muscle isolation. After isolating the target muscle activity, we recorded EMG and MUA, while an experimenter continuously monitored EMG for task compliance and proper muscle isolation. Each block consisted of well-isolated, repeated muscle contractions paced with a metronome for approximately 2.5 minutes (i.e. roughly 35 trials). The metronome cue occurred every 4 s (plus random jitter) and was presented simultaneously as an auditory beep and an onscreen flash of a gray patch (of duration 0.750 s). A photodiode marked the cue onset for referencing during offline analysis.

We excluded from further analysis trials where an EMG-producing muscle did not generate a recognizable signal or we observed extraneous muscle activity.

To rule out that neural activity was induced by posture or the metronome cues themselves, we repeated the experiment by having the participant sit in the same posture used during muscle contraction blocks, but passively listen and watch the metronome cues without moving.

### Sensorimotor Stability Experiment

To investigate stability of the sensorimotor map, we repeatedly characterized the sensorimotor activity and representation of the left extensor carpi radialis longus (ECRL) – a muscle that had a profuse representation among our arrays and could be reliably activated in an isolated manner without causing fatigue. We repeated the same metronome experiment described above across 12 sessions (approximately twice per week over the course of several months). During multiple sessions, we mapped ECR activity twice, approximately 2 hours apart. Day-to-day stability compared activity from one session to the next (roughly 4 days apart). Hour-to-hour stability compared activity collected at the beginning and end of a session (roughly 2 hours apart). We performed all analyses on trial-averaged PETHs from each block.

### Sensorimotor Map

We constructed (1) regional body maps (wrist: Fig. 2; fingers: Fig. S1); and (2) stability maps of the left ECRL to track somatotopy and individual channel activity over various timescales (Fig. 3A). For *regional body maps*, we labeled channels based on whether they were active for contractions of individual muscle groups (384 channels x 17 epochs x *m*, where *m* was number of isolated muscles per region, correcting for multiple comparisons using the False Discovery Rate, or FDR, method; see *Supplemental Information*^52-53^). For *stability maps*, we labeled channels according to the percentage of days that significant activity was registered (384 channels x 17 epochs x 12 days, corrected for multiple comparisons using FDR).

### Stability Definitions

#### Longitudinal Spatial Stability

We defined longitudinal spatial stability as frequency of sessions that a wrist contraction evoked activity on each electrode, and display these results as a heat map for each array (Fig. 4A). We estimated probabilities of survival (i.e. of being active for at least n sessions of 12) by counting the number of channels per area/ hemisphere active for at least n sessions, and converting these counts to probabilities by normalizing to the total number of active channels within that area/hemisphere. Shapes of probability functions were quantified by fitting exponential curves to each probability distribution of the form y=exp(-bn), with b estimating a survival rate.

#### Within-Channel Stability

We characterized unit-level stability of left ECRL representation in terms of *firing rate* and *firing dynamics. Firing Strength stability* is the normalized difference of z-scored PETH amplitudes. *Firing Dynamic stability* relates to correlation magnitude between aligned PETH waveforms, thus capturing more temporal features. Both metrics range from 0 to 1, higher values indicating greater stability. Computational details are available in *Supplementary Information*.

#### Stability Bounds

Although firing rate and firing dynamic stability are maximized at 1, this value is mathematical. We obtained practical upper limits for our data by bootstrapping on within-block data, reasoning that trials within the same block would be most stable. For each bootstrap experiment, we randomly selected a block of data, and sampled two sets of 40 trials with replacement from which to calculate a stability value. After completing 1000 such experiments, we computed the mean and 95% CI of stability statistics, as shown in Figs. 5A-D. Because such bounds were derived from within-block data, they also roughly estimate within-minute stabilities.

### Multi-Channel (Population) Stability and Decoding

The goal of our multi-channel analysis was to compare the population activity across electrodes implanted in common areas or hemispheres, in terms of both their similarity and their relationship to cued muscle activations in the left wrist extensor (ECRL). For averaging purposes on compliant trials only, we manually marked the EMGs for the beginning and end of the ECRL contractions to use as endpoints for a common time axis across trial, and then time-warped all EMG and neural signals to this axis by linear interpolation (150 points). Afterward, we averaged time-warped data by block.

In each block, we grouped neural channels into ensembles by brain area (motor/sensory), or hemisphere (contralateral/ ipsilateral), and analyzed them using principal component analysis (PCA). We wanted to assure that resulting PCs for each ensemble were uniform across all hour-to-hour and all day-to-day (session-to-session) blocks. To accomplish this, we first concatenated all blocks of ensemble-specific data for each time scale (hours or days), and then decomposed each ensemble-specific concatenation with a single PCA. Next, we applied the Procrustes algorithm^48,61^, which calculated a manifold alignment between consecutive trajectories using basic linear transformations (rotation, scaling, translation). It also computed a normalized squared error between the trajectories (increasing values ranging from 0 to 1 denoting greater instability), which we report.

To measure stability of the mapping between PC structure and EMG, we used a Wiener-filter approach, detailed in *Supplementary Information*.

### Statistical Analyses

We compared differences in longitudinal spatial stability by measuring the survival rates across time for channels of each area (motor, sensory) and hemisphere (contralateral, ipsilateral). Differences in the distribution of the number of sessions the channels between areas or hemispheres were assessed using nonparametric unpaired two-sample tests (Wilcoxon signed-rank test).

To compare within-channel firing rate and firing dynamic stability, we performed 2-way ANOVAs (1) with factors of brain area (motor, sensory) and timescale (hours, days), (2) with factors of laterality (contralateral, ipsilateral) and timescale (hours, days). We also relate to upper bounds by comparing mean values with 95% CIs of bounds.

We report all data as means ± 1 s.e. Effects were considered significant if p ≤ 0.05. All post-hoc analyses were done using two-tailed Tukey’s tests of Honestly Significant Difference (HSD).

## Data Availability

Data for this preprint is not publicly available at this time.

## ACKNOWLEDGMENTS

This research was supported by the following funding: the Defense Advanced Research Projects Agency (DARPA, Arlington, VA) Revolutionizing Prosthetics program (contract N6600110C4056) and Neurally-Enhanced Operations program (contract HR001120C0120); the National Institutes of Health (NIH) Eunice Kennedy Shriver National Institute of Child Health and Human Development (NICHD) T32HD741426 (RWN); and the National Institute of Neurological Disorders and Stroke (NIH-NINDS) NS088606 (TMT, DNC, NEC). The views, opinions, and/or findings expressed are those of the author(s) and should not be interpreted as representing the official views or policies of the Department of Defense or the U.S. Government. Development of experimental setup and support for regulatory submissions associated with this study were provided by a grant from the Alfred E. Mann Foundation. Software infrastructure and study preparation were developed with internal funding from Johns Hopkins University Applied Physics Laboratory and the Johns Hopkins School of Medicine.

We thank the participant for his participation in this and additional studies^36-37,50^. We gratefully acknowledge Drs. Kendra Cherry-Allen, Jennifer Keller, Vikram Chib, Robert Kambic, Agostina Casamento-Moran, and Charles E. Connor for helpful consultation. Additionally, we thank Dr. Peter Gorman, Dr. Cristina Sadowsky, and Dr. Philippines Cabahug and their staffs. This study was conducted under Investigational Device Exemption (IDE, 170010) by the Food and Drug Administration (FDA) for the purpose of evaluating bilateral sensory and motor capabilities of microelectrode array implants in people affected by tetraplegia. It is registered on clinicaltrials.gov as NCT03161067.

## AUTHOR CONTRIBUTIONS

Conceptualization: RWN, GLC, PAC;

Methodology: RWN, MAA, GLC, PAC

Validation: RWN, MAA, GLC, PAC

Formal Analysis, RWN analyzed data with input from MSF, DPM, GLC, PAC

Investigation: RWN, MAA, TMT, GLC, PAC

Resources: GLC, PAC

Data Curation: RWN

Writing – Original Draft: RWN, GLC

Writing ---Review & Editing: RWN, MAA, TMT, MSF, DPM, MCT, DNC, LEO, WSA, BAW, FVT, NEC, GLC, PAC

Visualization: RWN, BAW

Supervision: GLC, PAC

Project Administration: GLC, PAC

Funding Acquisition: FVT, GLC, PAC

## Notes

### Competing Interest Statement

The authors have declared no competing interest.

### Clinical Trial

NCT03161067

### Funding Statement

This research was supported by the following funding: the Defense Advanced Research Projects Agency (DARPA, Arlington) Revolutionizing Prosthetics program (contract N6600110C4056) and Neurally-Enhanced Operations program (contract HR001120C0120); the National Institutes of Health (NIH) Eunice Kennedy Shriver National Institute of Child Health and Human Development (NICHD) T32HD741426 (RWN); and the National Institute of Neurological Disorders and Stroke (NIH-NINDS) NS088606 (TMT, DNC, NEC). The views, opinions, and/or findings expressed are those of the author(s) and should not be interpreted as representing the official views or policies of the Department of Defense or the U.S. Government. Development of experimental setup and support for regulatory submissions associated with this study were provided by a grant from the Alfred E. Mann Foundation. Software infrastructure and study preparation were developed with internal funding from Johns Hopkins University Applied Physics Laboratory and the Johns Hopkins School of Medicine.

### Author Declarations

This study was conducted under an Investigational Device Exemption (IDE) by the FDA (G170010) and approved by the Johns Hopkins School of Medicine and NIWC Pacific Institutional Review Boards (IRBs)

