## Supplementary Information for "“Characteristics and stability of sensorimotor activity driven by isolated-muscle group activation in a human with tetraplegia”"

SUPPLEMENTAL METHODS

Map and stability analyses were primarily done on multiunit activity (MUA). During experiments, MUA signals were thresholded online (threshold level = -3.25dB relative to baseline activity measured at rest) and saved as a sequence of spike times (sampled at 30,000 Hz). Offline, we calculated spike times to firing rates by binning them at a frequency of 1 kHz to generate peristimulus time histograms (PETHs).

Most analyses were performed on firing rates represented by PETHs. However, as a side analysis we ran a spike sorting algorithm to study the relationship between stability and unit separability in our data (see section on *Cluster Analysis* section below).

**Active Channel Detection**

To determine body maps for the wrist muscles as in Figures 2C and 3A, we labeled the recording channels on our participant's arrays as active or inactive, based on the significance of their measured MUA. We determined which channels were active during contractions of wrist extensor and flexor muscles of each arm using a sliding window method (Fig. 1C).

For a given channel, *baseline activity* for each trial was calculated as the average firing rate (PETH amplitude) over a fixed window spanning -0.500s to -0.250s (relative to the EMG burst onset for that trial). *Response activity* for each trial was calculated at increments of 0.125s by sliding a window (length=0.250s, step width=0.125s) over the PETH from -0.250s to 2s and averaging the PETH amplitude within these windows. We detected MUA activations by comparing the average response firing rate for each window (17 total) to the mean baseline firing rate using separate two-sided statistical tests per channel. A paired t-test was used for channels with a normal distribution, and a two-sided Wilcoxson signed-rank test for non-normal distributions. We ascertained normality using Lilliefors tests. After correcting for multiple comparisons across channels and muscles using the false discovery rate (FDR) method at $q = 0.05$ [55-56], we labeled as active any channels having responses significantly different from baseline for at least 1 window. Channels that showed clear signs of muscle artifact were excluded from further analysis.

**Within-Channel Stability Calculations**

All stability calculations were done on PETHs for channels showing significant responses only.

*Firing rate stability:* Firing rate stability refers to the consistency of PETH amplitudes (normalized to baseline) across time, defined per channel as:

$$1 - \frac{|z_{t1} - z_{t2}|}{|z_{t1}| + |z_{t1}|}$$

where $z_{t1}$ and $z_{t2}$ refer to average PETH amplitudes in a window of -0.25 s to 0.25 s relative to the EMG burst, z-scored relative to baseline at measurement time points indexed by t1 and t2 (which may be separated by days or hours). We computed firing rate stability for each channel that was active during at least one endpoint of each time comparison (hour-to-hour, day-to-day).

*Firing dynamic stability:* Firing dynamic stability measures the similarity between the shape of the PETH waveforms at the two measurement times t1 and t2. It is defined per channel as the absolute value of the zero-lag cross-correlation between the z-scored PETHs within the aforementioned window about the EMG-burst. We calculated firing dynamic stability for each channel that was active during at least one endpoint of each time comparison (hour-to-hour, day-to-day).

Figure S2 shows example data that highlights changes in both within-channel stability types across consecutive sessions. We signify firing rate stability by $\Delta z$, and dynamic stability by $\rho$ .

**Cluster Analysis:**

Because previous studies have related channel stability to multiunit separability, i.e. single units may be more stable than multi-units[27], we compared whether the number of separable single units influenced our stability metrics. To do this, we sorted all channels into single-cluster (i.e. single unit) or multi-cluster (i.e. multiunit) groups using the unsupervised method `wave_clus`[59-60]. Using bootstrapping, we compared the relationships between brain area (M1, S1), multiunit separability (1 cluster only, or 2+ clusters), and firing dynamic stability.

**Decoder Calculations**

To measure the stability with which neural ensemble activity (expressed in PCs) encoded muscle activity during left wrist extensions, we applied a decoding approach similar to Gallego et al.[26]. Since we were interested in comparative stabilities between brain area and hemisphere, we trained decoders based on PETHs across 4 subsets (ensembles) of neural microelectrode data: all motor channels, sensory channels, contralateral channels (right hemisphere), and ipsilateral (left) channels. Before applying PCA, we culled out artifactual channels, and applied a square-root transform to all remaining PETHs. Outputs in all cases were the EMG envelopes from 6 muscles, comprising the wrist extensor, wrist flexor, and thumb adductor of each arm. Envelopes were filtered at 5 Hz, and for the purposes of model fitting, were normalized to the maximum amplitude of the left ECR envelope. The latter step served to mitigate variations in skin contact for electrodes across blocks.

The decoder model used was a Wiener filter, of the following form:

$$y[n] = \sum_{m=\mathrm{M1}}^{M2} A_m x[n-m]$$

Here, x[n] is the filter input, defined by the first 6 principal components across a channel ensemble at time point n; y is the filter output, defined as the vector of EMG envelopes over the above listed muscles at time n; and $A_m$ is the vector of mixing coefficients relating neural input to muscular output. Index m denotes the number of time instants (or “taps”) factored into the model before ($m > 0$), or after ($m < 0$) the instant of EMG measurement. We downsampled all data at 100 Hz, and considered inputs comprising (1) motor array channels only, (2) sensory array channels only, and (3) both array types. To model the EMG output at time n, we included the previous 100 ms (10 timesteps) of neural data for motor-input models ($M1= 0$, $M2 = 10$), the following 100 ms (10 timesteps) for sensory-input models ($M1 = -10$, $M2 = 0$), and a symmetric window of 100 ms ($M1 = -5$, $M2 = 5$) for mixed-input models. These values comport with those used in Gallego et al.[26], and agree with neural latency estimates from our data (Fig S3).

For each pair of consecutive data blocks (spaced by hours or by days/sessions), we designated one block as a training set, and the other as a test set. We estimated the Wiener filter using the PCs on the first data set of the temporal pair, and cross validated this filter against the neural activity and EMG of the test data set. Stability was measured as the correlation between actual and predicted EMG of the left wrist with $R^2$ (see Fig 5E-F). We then interchanged the training and test sets and repeated this process. Both $R^2$ values were then averaged to yield an overall goodness of fit for the consecutive-timepoint comparison, and the overall process was repeated across all pairs of consecutive times. This process, when carried out for all possible neural data ensembles (motor/sensory or contralateral/ipsilateral, yielded the distribution of $R^2$ values presented in Fig 5G-H.

**Multiunit Latency**

We estimated latency distributions for MUA evoked by ECR contractions, in order to verify assumptions of our Wiener-filter decoding model (shown in Fig S3). First, for each active channel, we averaged the EMG-referenced PETHs over trials. Then we estimated latencies based from times when mean PETHs crossed a threshold (the temporal mean of the baseline window plus or minus a multiple of its temporal standard deviation) and remained above or below for at least 0.050 s consecutively. For left ECR, we set this multiple to 3.5 times baseline s.d. This multiple was approximately equal to the Bonferroni-corrected threshold labeling a channel active (0.05 significance level), and is comparable to other published latency thresholds[54-58]. In final analysis, we rejected as outliers any data outside the median + 1.5 times the interquartile ratio (corresponding to average retentions of 88.4% of motor and 82.8% of sensory values).

Latencies for the left ECR were approximately Gaussian distributed for both motor and sensory activity and centered relative to EMG burst onset (Fig S3). Average motor latencies preceded sensory latencies (mean motor latency = $x_M$ = 0.0017 +/- 0.0037 s relative to EMG onset; mean sensory latency $x_S$ = 0.069 +/- 0.0022 s relative to EMG onset). Moreover, a subset of sensory latencies occurred prior to 30 ms after the initial EMG burst (denoted by inclusion in the pink shaded region of Fig S3), preceding what we expected for a normal cortico-muscular delay.

SUPPLEMENTAL FIGURES

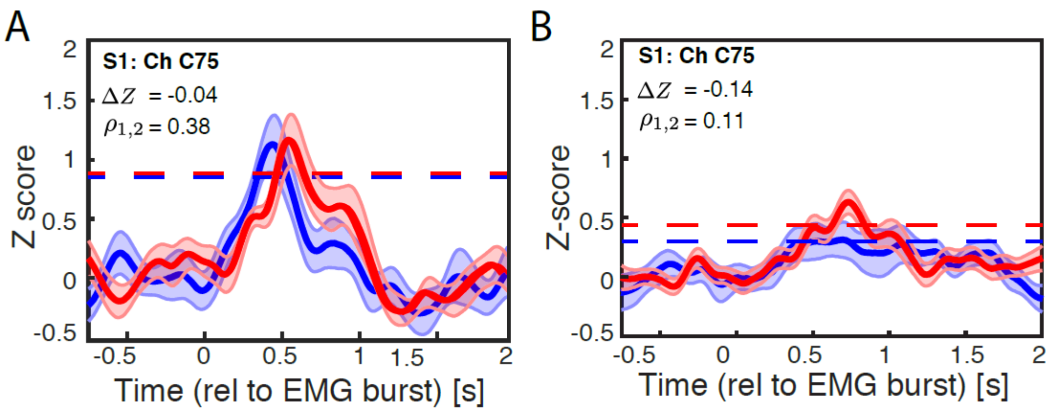


**Figure S1: Declining firing--rate and firing--dynamic stability over time for left ECR-related activity, on a representative channel (Pedestal C sensory array, Channel 75).**

(A-B) Red and blue curves denote z-scored PETHs from each endpoint of the time comparison (means +/- 1 s.e. from bootstrapping). Firing strength stability is reported as a change in average z-scored PETH over an interval [-0.25, 0.25], and is represented numerically with the variable $\Delta Z$, and graphically for each time point as a horizontal dashed line. Firing dynamic stability is given as magnitude of cross correlation (zero-lag) between waveforms in the same interval, and denoted numerically with the variable $\rho_{1,2}$.

A) Representative firing rate across two measurements on the time scale of minutes.

B) Representative firing rate across two measurements on the time scale of days. Note The overall increase in $\Delta Z$, and decrease in $\rho_{1,2}$ that are shown in panel B reflect that these waveforms become less similar across time points, and thus the within-channel firing patterns destabilize.

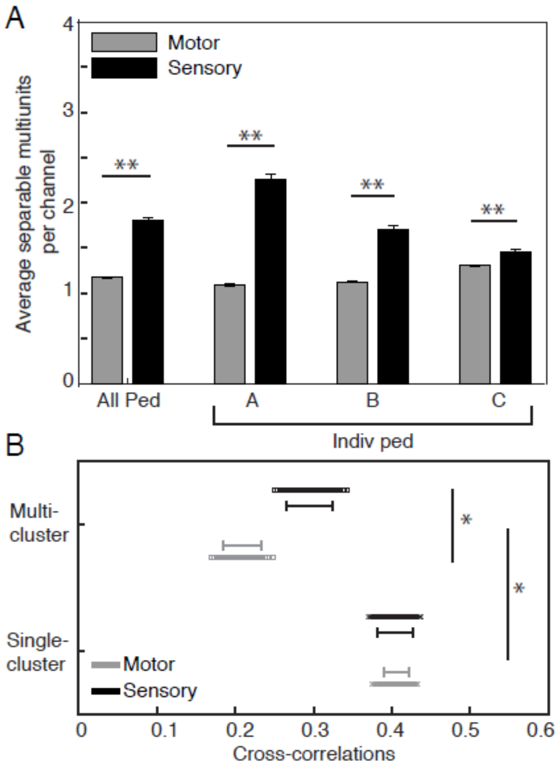


**Figure S2: Influence of multiunit separability on within-channel stability.**

(A-B) Error bars are +/- 1 s.e. Annotations (*) and (**) denote significance levels of 0.05 and 0.01, respectively.

A) Number of separable units for each brain region over pedestals, as determined by wave_clus method. Data is shown for aggregate (left), and individual pedestals.

B) Bootstrapped distributions of firing dynamic stability, expressed as cross-correlation in [-0.25, 0.25] s response window. Higher stability is denoted by higher values. Format is as in panel B. Results suggest that for channels with multiple separable clusters, stability in the sensory cortex is higher than for the motor cortex.

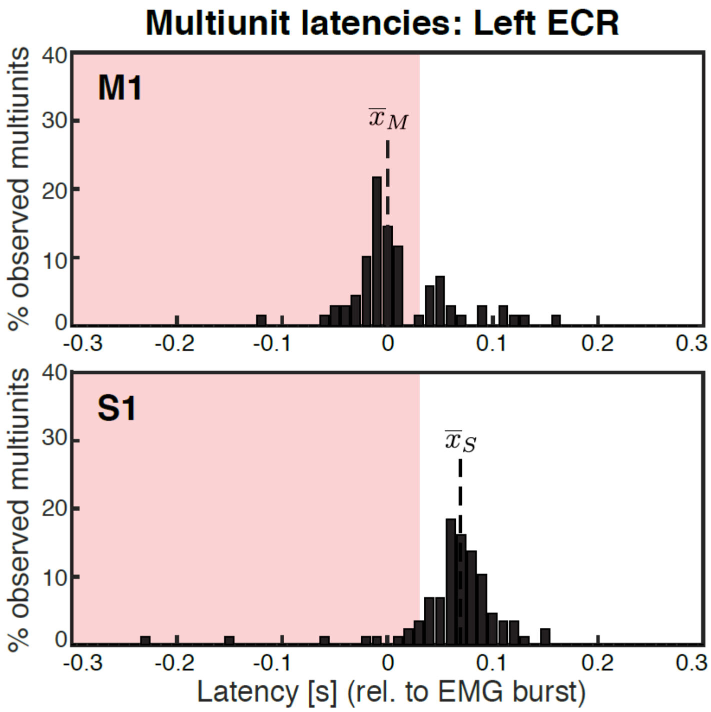


**Figure S3: Neural latency distributions on motor (top row) and sensory (bottom) channels responding to left ECR contractions.**

The distribution of latencies in the motor (top panel) and sensory arrays (bottom) are shown, relative to EMG burst onsets (t = 0). Vertical dashed lines labeled $\overline{x_M}$ and $\overline{x_S}$ are mean motor and sensory latencies, respectively. Pink regions highlight potential efference activity at latencies below 30 ms, Virtually all counts are from the pedestals contralateral to the side of the body of the muscle. Latencies outside of the range of the median +/- 1.5 times the interquartile range were excluded, per a conventional definition of outliers for non-parametric distributions. This amounted to retentions of 88.4% of motor and 82.8% of sensory latency values.
